# Effectiveness of a third dose of BNT162b2 or mRNA-1273 vaccine for preventing post-vaccination COVID-19 infection: an observational study

**DOI:** 10.1101/2021.11.29.21266777

**Authors:** Aditya Sharma, Gina Oda, Mark Holodniy

## Abstract

**Background:** BNT162b2 and mRNA-1273 vaccines are highly protective against COVID-19. Concern about waning immunity and reduced effectiveness against SARS-COV-2 variants led to use of a third dose six months after completing the primary series. We used data from the Veterans Health Administration to evaluate the effectiveness of a third dose of BNT162b2 or mRNA-1273 compared to the primary series in preventing post-vaccination COVID-19.

**Methods:** During January 1 - November 25, 2021, third dose recipients were matched (1:1) to demographically similar controls who did not receive a third dose. Eligible participants had completed the primary series at least six months (180 days) before recruitment date and had received at least one SARS-CoV-2 PCR test. Long-term care residents were excluded. Primary outcomes were documented SARS-CoV-2 infection and COVID-19 hospitalization. Effectiveness was estimated as 1-incidence rate ratio.

**Findings:** Following matching, the BNT162b2 group included 74,032 pairs and the mRNA-1273 group included 55,098 pairs. In BNT162b2 and mRNA-1273 groups, median age was 72 (interquartile range [IQR]: 64-75) and 72 (IQR: 66-77) years, 69,426 (93.8%) and 52,241 (94.8%) were male, and 43,039 (58.1%) and 37,218 (67.5%) were non-Hispanic White, respectively. Effectiveness of a third dose of BNT162b2 or mRNA-1273 compared to the primary series was 45.7% (95% confidence interval [CI]: 37.9-52.5%) and 46.6% (95% CI: 36.4-55.3%) for documented SARS-CoV-2 infection and 44.8% (95% CI: 26.6-58.4%) and 50.0% (95% CI: 26.2-66.1%) for COVID-19 hospitalization, respectively.

**Interpretation:** A third dose of BNT162b2 or mRNA-1273 is moderately effective against post-vaccination COVID-19 infection compared to the primary series.

**Funding:** None.

## Background

Messenger RNA (mRNA) based vaccines have demonstrated significant protection against COVID-19 compared to unvaccinated persons in clinical as well as in observational studies^1-4^. The spread of the SARS-CoV-2 delta variant, resurgence of COVID-19 infections, and concern about waning antibody levels among vaccinated persons led US Food and Drug Administration (FDA) to authorize a third dose of BNT162b2 (Pfizer-BioNTech) and mRNA-1273 (Moderna) 6 months after completing the primary series for vaccinated persons at high risk of severe disease or exposure to COVID-19^5,6^. Soon after, several states expanded eligibility of booster doses to all adults as part of a continued effort to control COVID-19^7-9^. On November 19, 2021 FDA expanded eligibility for booster vaccine doses to all individuals 18 years and older after completing the primary vaccine series^10^. By November 22, 2021, US Centers for Disease Control and Prevention (CDC) estimated that more than 36 million individuals and at least 41% of fully vaccinated persons aged 65 and older in the United States had received an additional dose of vaccine^11^.

Despite the rapid uptake of vaccine doses after completing the primary series and the public health importance of post-vaccination infection, the effectiveness of a third dose of BNT162b2 or mRNA-1273 has not been sufficiently studied in a real-world context. Most studies describe substantial rises in antibody titers within days of administration of a third dose^12,13^, though it is unclear whether this rise is accompanied by reduced risk of clinical disease. Observational studies in Israel demonstrated effectiveness of a third dose of BNT162b2 in preventing post-vaccination COVID-19 compared to persons who only completed the primary series^14,15^. However, the effectiveness of a third dose of either mRNA-based vaccine has yet to be described in the United States.

We leveraged electronic health records from the Veterans Health Administration (VHA), the largest integrated health system in the United States, to estimate the effectiveness of a third dose of BNT162b2 and mRNA-1273 vaccine in preventing post-vaccination COVID-19 infection.

## Methods

We followed a methodological approach of emulating a target trial of the effects of a third dose of vaccine in a population who had completed the primary series at least 6 months (180 days) before recruitment^16^; this approach is similar to that used in other recent observational studies^15,17,18^.

VHA is the largest integrated health system in the United States, providing healthcare services at 1,293 facilities^19^. Individual-level clinical records are parsed and imported into the VHA Corporate Data Warehouse, which is used to conduct observational studies as well as to monitor multi-level operations. VHA implemented vaccination for COVID-19 beginning in December 2020. The study period was January 1, 2021 to November 25, 2021. For study inclusion, an individual had to have received the second dose of BNT162b2 or mRNA-1273 at least 6 months (180 days) before recruitment. Additionally, to focus on active users, individuals needed to have received at least one SARS-CoV-2 PCR test after full vaccination. Persons living in long-term care facilities were excluded.

### Procedures

For each vaccine, the target trial approach for this study was designed to compare two treatment strategies: completion of the primary series and no third dose of vaccine versus completion of the primary series and administration of a third dose of vaccine. Individuals who received the third dose of vaccine on a particular day during the study period were matched to persons who had completed the primary series but had not received the third dose. Persons who did not receive a third dose could be recruited in the third dose group if the third dose was given during the follow up period. Individuals were matched on multiple demographic and clinical covariates: age, sex, race/ethnicity, calendar month of completing the primary series, comorbidities (summarized by the Charlson comorbidity index), number of SARS-CoV-2 PCR tests received and history of positive SARS-CoV-2 PCR test in the 90 days before recruitment, and state of residence.

### Outcomes

Two post-vaccination infections were considered as outcomes: documented SARS-CoV-2 infection (defined as a positive SARS-CoV-2 PCR test) and COVID-19 hospitalization (defined as documented SARS-CoV-2 infection 14 days before or 3 days after admission to an inpatient unit in an acute care facility). For each outcome and vaccine, matched pairs were followed from recruitment until the earliest date of outcome, death, or end of the study period. Follow up was also halted if the matched non-recipient received the third dose. Outcomes were assessed in the period after administration of a third dose.

### Statistical analysis

Incidence rate ratios and corresponding 95% confidence intervals (CI) for documented SARS-CoV-2 infection and COVID-19 hospitalization were calculated; estimates were also stratified by age group and Charlson comorbidity score. Effectiveness of receiving a third dose of vaccine was estimated as 1-incidence rate ratio. The Kaplan-Meier estimator was used to calculate cumulative incidence of documented SARS-CoV-2 infection and COVID-19 hospitalization stratified by having received a third dose of vaccine during a 90-day period. All analyses were performed in R version 4.10^20^.

This project was approved by the Stanford University Institutional Review Board (Protocol ID 47191, “Public Health Surveillance in the Department of Veterans Affairs”) and written informed consent was waived.

## Results

During January 1 - November 25, 2021, 220,447 individuals who received BNT162b2 and 218,762 persons vaccinated with mRNA-1273 met eligibility criteria. Among these, 79,458 (36.0%) and 59,018 (27.0%) individuals received a third dose of BNT162b2 or mRNA-1273, respectively. After matching, the BNT162b2 group consisted of 74,032 pairs and the mRNA-1273 group included 55,098 pairs of vaccinated individuals.

Baseline demographic characteristics were similar across vaccine groups and pairs (Table 1). Among subgroups of persons vaccinated with BNT162b2, median age was 72 (interquartile range [IQR]: 64-75) years, 69,426 (93.8%) were male, 43,039 (58.1%) were non-Hispanic White, and 40,272 (54.4%) had a Charlson comorbidity index of ≥ 5. Similarly, across pairs of persons vaccinated with mRNA-1273, median age was 72 (IQR: 66-77) years, 52,241 (94.8%) were male, 37,218 (67.5%) were non-Hispanic White, and 30,750 (55.8%) had a Charlson comorbidity index of ≥ 5. Less than 2% of pairs in both vaccine groups had a positive SARS-CoV-2 PCR test and the majority did not undergo SARS-CoV-2 testing in the 90 days before start of follow up.

**Table 1.** Characteristics and outcomes of study participants in BNT162b2 and mRNA-1273 groups stratified by receipt of third dose of vaccine.

| Characteristic | BNT162b2 |  | mRNA-1273 |  |
| --- | --- | --- | --- | --- |
|  | Primary series<br>N = 74,032 | Third dose<br>N = 74,032 | Primary series<br>N = 55,098 | Third dose<br>N = 55,098 |
| Age (years)<sup>1</sup> | 72 (64, 75) | 72 (65, 75) | 72 (66, 76) | 72 (66, 76) |
| Age group (years) |  |  |  |  |
| 18-44 | 1,755 (2.4%) | 1,671 (2.3%) | 971 (1.8%) | 920 (1.7%) |
| 45-64 | 16,795 (22.7%) | 15,716 (21.2%) | 10,583 (19.2%) | 10,144 (18.4%) |
| 65+ | 55,482 (74.9%) | 56,645 (76.5%) | 43,544 (79.0%) | 44,034 (79.9%) |
| Sex |  |  |  |  |
| Female | 4,606 (6.2%) | 4,606 (6.2%) | 2,857 (5.2%) | 2,857 (5.2%) |
| Male | 69,426 (93.8%) | 69,426 (93.8%) | 52,241 (94.8%) | 52,241 (94.8%) |
| Race/Ethnicity |  |  |  |  |
| Hispanic or Latino | 5,148 (7.0%) | 5,148 (7.0%) | 4,522 (8.2%) | 4,522 (8.2%) |
| Non-Hispanic Black | 20,453 (27.6%) | 20,453 (27.6%) | 9,270 (16.8%) | 9,270 (16.8%) |
| Non-Hispanic White | 43,039 (58.1%) | 43,039 (58.1%) | 37,218 (67.5%) | 37,218 (67.5%) |
| Other | 5,392 (7.3%) | 5,392 (7.3%) | 4,088 (7.4%) | 4,088 (7.4%) |
| Charlson comorbidity index |  |  |  |  |
| 0 | 3,419 (4.6%) | 3,419 (4.6%) | 1,838 (3.3%) | 1,838 (3.3%) |
| 1-2 | 12,629 (17.1%) | 12,629 (17.1%) | 8,998 (16.3%) | 8,998 (16.3%) |
| 3-4 | 17,712 (23.9%) | 17,712 (23.9%) | 13,512 (24.5%) | 13,512 (24.5%) |
| 5+ | 40,272 (54.4%) | 40,272 (54.4%) | 30,750 (55.8%) | 30,750 (55.8%) |
| Number of Previous SARS-CoV-2 PCR Tests<sup>2</sup> |  |  |  |  |
| 0 | 38,879 (52.5%) | 38,879 (52.5%) | 27,996 (50.8%) | 27,996 (50.8%) |
| 1-2 | 32,807 (44.3%) | 32,807 (44.3%) | 25,496 (46.3%) | 25,496 (46.3%) |
| 3-4 | 1,676 (2.3%) | 1,676 (2.3%) | 1,064 (1.9%) | 1,064 (1.9%) |
| 5+ | 670 (0.9%) | 670 (0.9%) | 542 (1.0%) | 542 (1.0%) |
| Previous documented SARS-CoV-2 infection<sup>2</sup> | 1,085 (1.5%) | 1,085 (1.5%) | 601 (1.1%) | 601 (1.1%) |
| Documented SARS-CoV-2 Infection | 604 (0.8%) | 328 (0.4%) | 356 (0.6%) | 190 (0.3%) |
| Follow-up time (days)<sup>1</sup> | 30 (14, 44) | 30 (14, 44) | 16 (8, 25) | 16 (8, 25) |
| COVID-19 Hospitalization | 134 (0.2%) | 74 (0.1%) | 76 (0.1%) | 38 (0.1%) |
| Follow-up time (days)<sup>1</sup> | 30 (14, 44) | 30 (14, 44) | 16 (8, 25) | 16 (8, 25) |

Numbers describe n (%) unless otherwise noted.
<sup>1</sup>Median (IQR)
<sup>2</sup>90 days before start of follow up

In the BNT162b2 group, documented SARS-CoV-2 occurred in 328 (0.4%) of third dose recipients and 604 (0.8%) of non-recipients; COVID-19 hospitalization occurred in 74 (0.1%) and 134 (0.2%), respectively. Median follow up time was 30 (IQR: 14 - 44) days for both documented SARS-CoV-2 infection and COVID-19 hospitalization. In the mRNA-1273 group, documented SARS-CoV-2 occurred in 190 (0.3%) of persons who received a third dose compared to 356 (0.6%) non-recipients; COVID-19 hospitalization occurred in 38 (0.1%) and 76 (0.1%), respectively. Median follow up time for both outcomes was 16 (IQR: 8 - 25) days.

Estimated effectiveness of a third dose of BNT162b2 was 45.7% (95% confidence interval [CI]: 37.9 - 52.5%, p < 0.001) for documented SARS-CoV-2 infection and 44.8% (95% CI: 26.6 - 58.4%, p < 0.001) for COVID-19 hospitalization (Table 2). Estimated effectiveness of a third dose of mRNA-1273 was 46.6% (95% CI: 36.4 - 55.3%, p < 0.001) for documented SARS-CoV-2 infection and 50.0% (95% CI: 26.2 - 66.1%, p < 0.001) for COVID-19 hospitalization. In a stratified analysis, effectiveness of a third dose against documented SARS-CoV-2 infection was similar across age groups, but marginally higher in individuals with lower comorbidity scores (Table 3). Effectiveness of a third dose against COVID-19 hospitalization was similar across comorbidity scores but did not appear to be substantial in older persons.

**Table 2.**
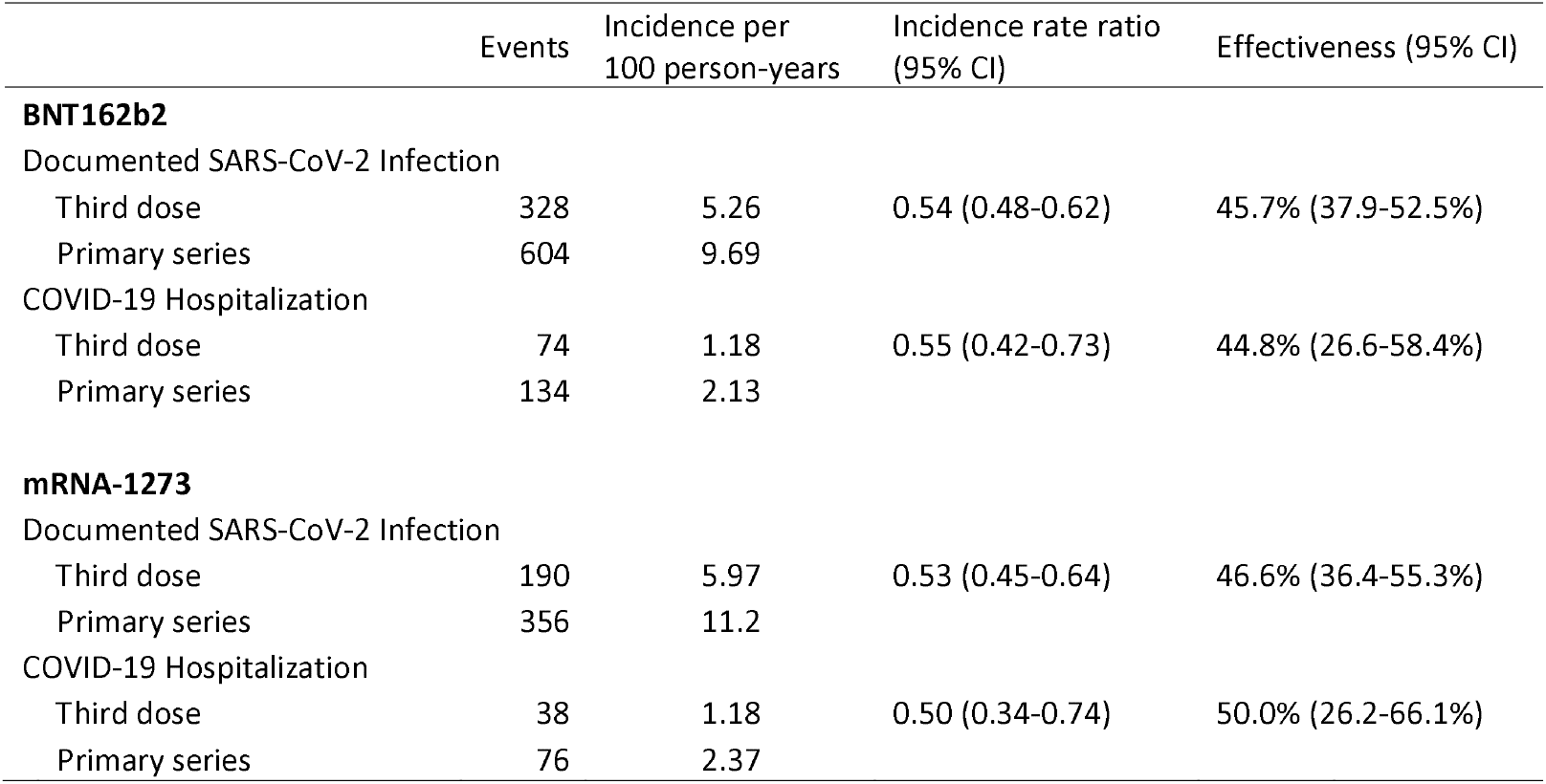
Estimated effectiveness of third dose of BNT162b2 or mRNA-1273 vaccines compared to the primary series.

**Table 3.** Estimated effectivess of third dose of BNT162b2 or mRNA-1273 compared to primary series stratified by selected age groups and Charlson comorbidity scores.

|  | **Documented SARS-CoV-2 infection** |  | **COVID-19 hospitalization** |  |
| --- | --- | --- | --- | --- |
|  | Incidence rate ratio<br>(95% CI) | Effectiveness<br>(95% CI) | Incidence rate ratio<br>(95% CI) | Effectiveness<br>(95% CI) |
| **BNT162b2** |  |  |  |  |
| Age group |  |  |  |  |
| <65 years | 0.55 (0.47-0.63) | 45.4% (36.5-53.1%) | 0.53 (0.39-0.71) | 47.3% (28.9-60.9%) |
| 65+ years | 0.53 (0.40-0.71) | 46.9% (28.9-60.3%) | 0.73 (0.30-1.79) | 26.8% (-78.9-70.1%) |
| Charlson comorbidity score |  |  |  |  |
| <5 | 0.45 (0.36-0.55) | 55.5% (45.3-63.8%) | 0.44 (0.23-0.82) | 56.3% (18.0-76.7%) |
| 5+ | 0.63 (0.53-0.76) | 36.5% (24.2-46.9%) | 0.59 (0.43-0.81) | 41.2% (19.1-57.2%) |
| **mRNA-1273** |  |  |  |  |
| Age group |  |  |  |  |
| <65 years | 0.54 (0.45-0.65) | 46.3% (35.1-55.5%) | 0.50 (0.33-0.74) | 50.4% (25.6-66.9%) |
| 65+ years | 0.49 (0.30-0.81) | 50.6% (18.7-70.0%) | 0.52 (0.13-2.04) | 48.2% (-104.4-86.9%) |
| Charlson comorbidity score |  |  |  |  |
| <5 | 0.51 (0.39-0.68) | 48.6% (31.8-61.2%) | 0.54 (0.21-0.98) | 46.2% (2.1-78.5%) |
| 5+ | 0.55 (0.44-0.68) | 45.3% (31.5-56.4%) | 0.49 (0.32-0.76) | 50.8% (24.4-68.0%) |

Among individuals vaccinated with BNT162b2, the 90-day cumulative incidence of documented SARS-COV-2 infection and COVID-19 hospitalization was 1.0% (95% CI: 0.8 - 1.3%) and 0.3% (95% CI: 0.1 - 0.4%) in recipients of a third dose compared to 1.9% (95% CI: 1.5 - 2.3%) and 0.5% (95% CI: 0.2 - 0.8%) in persons who did not receive a third dose, respectively (Figure 1). Among persons who received mRNA-1273, the 90-day cumulative incidence of documented SARS-COV-2 infection and COVID-19 hospitalization was 1.4% (95% CI: 1.0 - 1.8%) and 0.2% (95% CI: 0.1 - 0.3%) in recipients of a third dose compared to 2.6% (95% CI: 1.9 - 3.2%) and 0.5% (95% CI: 0.3 - 0.6%) in persons who did not receive a third dose, respectively (Figure 2). For both vaccines, differences in cumulative incidence became noticeable within the first 5 days after administration of the third dose for documented SARS-CoV-2 infection and 10 days for COVID-19 hospitalization.

**Figure 1.**
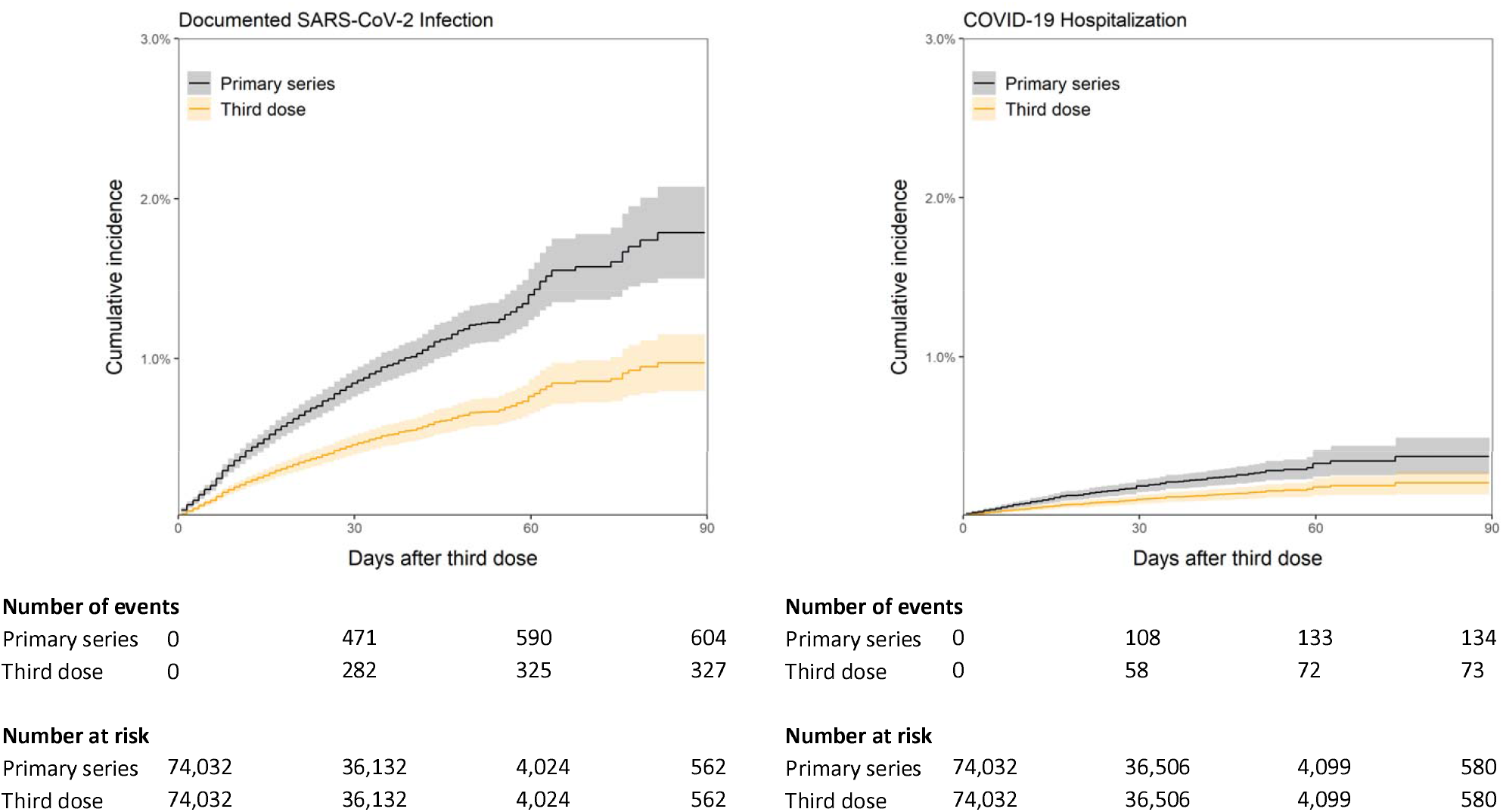
Cumulative incidence of documented SARS-CoV-2 infection and COVID-19 hospitalization in individuals who received a third dose of BNT162b2 compared to those who completed the primary series.

**Figure 2.**
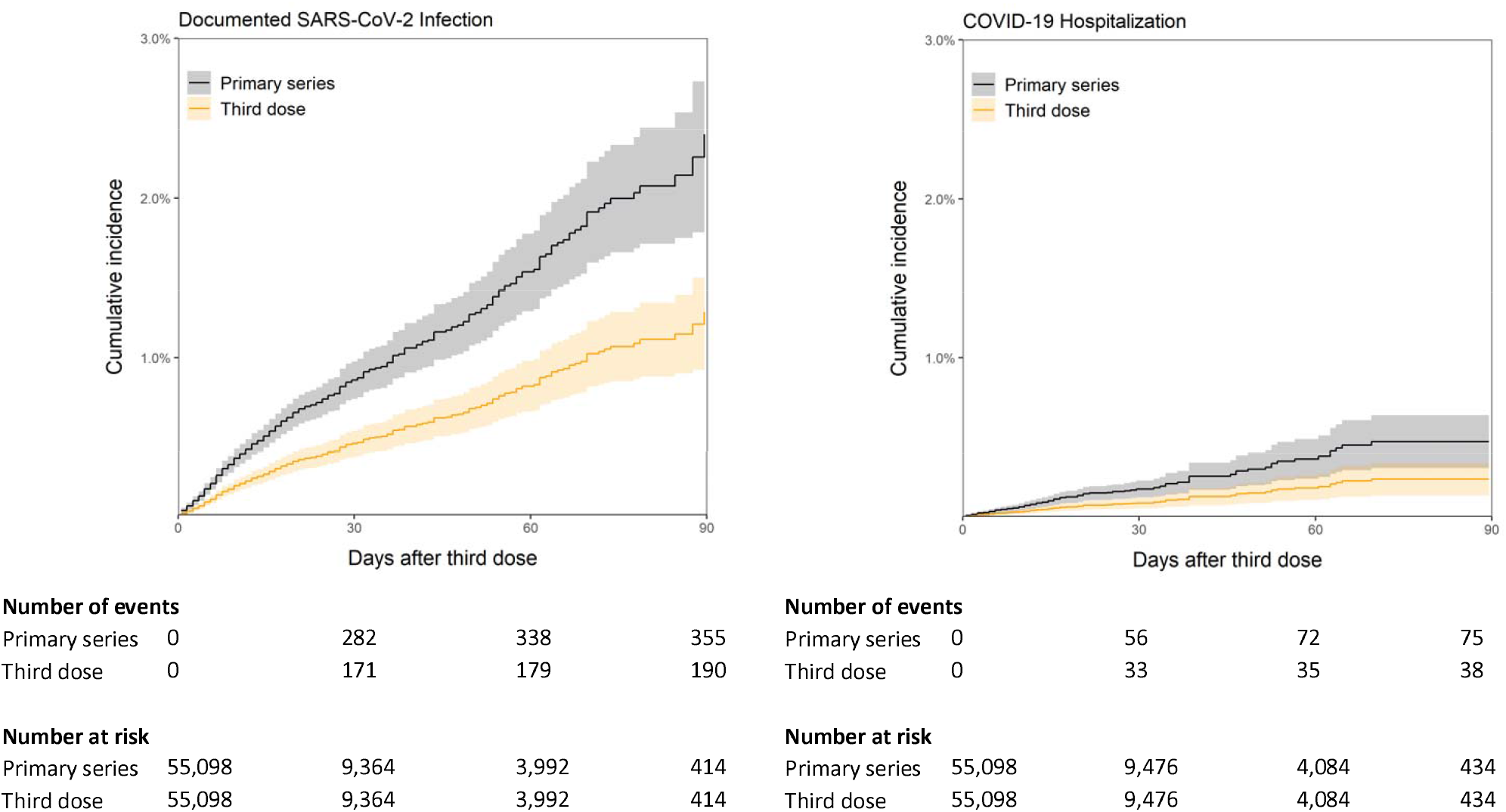
Cumulative incidence of documented SARS-CoV-2 infection and COVID-19 hospitalization in individuals who received a third dose of mRNA-1273 compared to those who completed the primary series.

## Discussion

We estimated the effectiveness of a third dose of BNT162b2 or mRNA-1273 vaccine in persons who completed the primary series and received care in the largest healthcare system in the United States. A third dose of BNT162b2 administered at least 6 months after completing the primary series was found to be 45.7% effective in preventing documented SARS-CoV-2 infection and 44.8% in preventing COVID-19 hospitalization. Additionally, a third dose of mRNA-1273 was 46.6% effective in preventing documented SARS-CoV-2 infection and 50.0% in preventing COVID-19 hospitalization. Our findings demonstrate moderate reductions in post-vaccination infections in persons who receive a third dose of vaccine 6 months after completing the primary series of BNT162b2 or mRNA-1273; improvements in documented SARS-CoV-2 infection and COVID-19 hospitalization occur within 5 and 10 days of administration of a third dose, respectively.

Our findings demonstrate lower effectiveness of a third dose of BNT162b2 compared to a similar study in Israel by Barda et al^15^. Another study in Israel measured high effectiveness of a third dose compared to the primary series, though estimates were not adjusted for comorbidities^21^. Observed differences in our estimates and those in Barda et al might be explained by variation in COVID-19 incidence during the study periods as well as heterogeneity in the implementation of and adherence to non-pharmaceutical interventions to limit spread; it is likely that a third dose of vaccine would appear more effective in settings with higher intensity of COVID-19 transmission. Additionally, the study population in Barda et al consisted of younger-aged individuals with few comorbidities, while our study population generally includes older persons with higher comorbidity scores who had at least one SARS-CoV-2 test after completing the primary series. Therefore, our study is more representative of individuals with higher baseline risk of COVID-19 infection.

We observed that both mild and severe post-vaccination infections (approximated in our study as documented SARS-CoV-2 infection and COVID-19 hospitalization, respectively) occur less frequently in persons who received the third dose of vaccine, demonstrating a modest individual-level health benefit. Reduced frequency of outcomes in persons who received a third dose of BNT162b2 or mRNA-1273 might be explained by restored antibody levels that would otherwise have been reduced 6 months after completing the primary series. A study from Israel demonstrated that fully vaccinated individuals carry lower viral loads compared to unvaccinated persons; this effect was observed to disappear 6 months after vaccination but is restored after administration of a third dose of BNT162b2^22^. However, it is unknown whether the restored effect due to a third dose is temporary, and if administration of additional doses at regular intervals would be necessary to maintain protection from possible infection. Additionally, our observations including those in the stratified analysis might be explained by differences in exposures: those sufficiently concerned about post-vaccination infection to seek an additional vaccine dose might also choose less risky behaviors than non-recipients; therefore, differences in general preventive behaviors between third dose recipients and non-recipients might confound the frequency of observed outcomes. Older persons or those with higher number of comorbidities might not engage in as many social activities compared to younger, healthier individuals, and consequently might not experience comparable transmission risk. Our study also suggests longitudinal protection of the primary series of mRNA-based vaccines against severe COVID-19, with less than 1% of individuals in this subgroup experiencing COVID-19 hospitalization 90 days after start of follow up, which in turn is at least 6 months after full vaccination.

There has been considerable debate regarding the public health benefit of administering third doses of mRNA-based vaccines in persons who completed the primary series in comparison to a strategy of encouraging uptake of vaccine doses in unvaccinated persons. Though we did observe a modest improvement in post-vaccination infection with administration of a third dose, it is unknown whether this improvement also translates to reduced frequency of secondary infections among contacts. A large observational study in the United Kingdom demonstrated that vaccinated persons carry similar peak viral loads as unvaccinated persons but for a shorter duration^23^. Additionally, the secondary attack rate among household contacts did not differ by vaccination status of the index case, and transmission of delta variant was observed to occur among fully vaccinated index case-contact pairs. At the population level, it is likely that further suppression of COVID-19 transmission will occur as unvaccinated subgroups consisting of younger and more active individuals become fully vaccinated, while booster vaccine doses will remain relevant for preventing severe infection among high-risk individuals.

Our findings are subject to several limitations. Clinical records for patients who received care in facilities external to VHA might not be available in VHA databases unless these services were ordered by VHA providers and paid for by VHA; therefore, these testing episodes and outcomes would be missed in our analysis. Although VHA issued national testing guidelines, differences across VHA facilities in testing assays and local policies or approaches to testing may contribute to variability in detection of vaccine breakthrough events; some events might have been missed or misclassified. Our study population consists of predominantly older, male persons; therefore, our results might not be generalizable to the larger US population. Though we sought to control for health-seeking behavior as well as demographic and clinical covariates associated with COVID-19, unmeasured confounders might affect our findings. To reduce residual confounding, we excluded long term care residents; our findings might not be applicable to this subgroup. Though we observed modest effectiveness of a third dose of vaccine in reducing post-vaccination infection, we were unable to assess associations with broader epidemiological outcomes such as reduced transmission among contacts. Given the observational nature of this study, data describing additional biomarkers, timing of exposures, symptoms, and the specific variants occurring in vaccine breakthrough events were unavailable; therefore, we were unable to assess the importance of these factors with post-vaccination infection.

In summary, we observed moderate effectiveness of a third dose of BNT162b2 or mRNA-1273 administered at least 6 months after completing the primary series. Compared to persons who completed the primary series, those who received a third dose experienced fewer episodes of documented SARS-CoV-2 infection and severe COVID-19 disease requiring hospitalization. Clinicians and public health administrators should consider these findings in the broader context of patient- and population-level efforts to combat COVID-19.

## Data Availability

Due to US Department of Veterans Affairs (VA) regulations, the analytic datasets used for this study are not permitted to leave the VA firewall without a Data Use Agreement. This limitation is consistent with other studies based on VA data.

## Contributions

Conceptualization: AS, GO, MH

Data curation: AS

Formal analysis: AS

Methodology: AS, GO, MH

Project administration: AS, GO, MH

Resources: AS, GO, MH

Software: AS

Supervision: MH

Visualization: AS

Writing (original draft): AS

Writing (review & editing): AS, GO, MH

